# Variant calling pipelines benchmark for whole exome sequencing in clinical context

**DOI:** 10.1101/2024.10.18.24315708

**Authors:** Johanna Stepanian, Diego Saldaña, Daniel Mahecha, Laura Mazabel, Jorge Diaz

## Abstract

1

**Introduction:** Whole exome sequencing (WES) has become a more accessible diagnostic tool in clinical genetic context, leading to the debate of the most accurate and effective bioinformatic pipeline solutions to evaluate variants that explain diseases.

**Objective:** This study aimed to evaluate twenty-four pipelines in two samples comparing accuracy, time and computing efficiency. We also contrasted the results based on regions in two of the most common capture kits.

**Materials and methods:** We used two accessions of NA12872 whole exome sequencing to contrast four different free access software for mapping using hg38 reference genome, then we used six different software alternatives for variant calling process. Finally, differences in computational resources and efficacy were evaluated.

**Results:** Our results showed that the most accurate and fastest pipeline is BWA with Strelka for SNVs detection, and differences in the use of resources and efficacy were proven.

**Conclusions:** BWA and Strelka are the most accurate and fastest for detecting SNVs in clinical exomes. Significant differences in efficiency and resource usage exist among the workflows evaluated. These findings aid in selecting the best methods for clinical contexts.

## 2 Introduction

Since the introduction of DNA sequencing by Fred Sanger in 1977 and the subsequent development of Next-Generation Sequencing (NGS), the data resulting from the extensive application of these technologies in diverse fields have spurred the development of numerous algorithms to analyze vast datasets. Decreasing NGS costs has facilitated the utilization of these tools not only in research but also for clinical diagnosis, enabling the analysis of genetic variability in human populations and the identification of variants related to monogenic conditions and complex diseases (1). These advancements have optimized both the time and precision of results, contributing to the advancement of genomic medicine (2–6).

Millions of individual exomes have been sequenced worldwide (7,8) with the primary objective of exploring variants encoded within these genomic sequences through a process known as variant calling. This process comprises two essential steps: mapping the sequenced DNA against a reference genome and identifying the set of variants (differences in sequences when compared with the reference) that cannot be attributed to sequencing errors. The accuracy and reliability of variant calling are influenced by various factors throughout this process, including the quality of the sequences, the reference genome, and the software employed (9).

The entire process of detecting genetic variants is a crucial procedure with a significant impact over the interpretation of genomic data and its application in medical diagnosis. Consequently, multiple recommendations have been issued regarding the need for caution in the processing and interpretation of genetic tests due to the complexity of genomic information and the health risks associated with these tools, given the probabilistic nature of available analysis algorithms (10). Therefore, the evaluation and comparison of various bioinformatic tools and methods for variant mapping and calling are imperative for the best analysis of genomic data.

Currently, genomic variants such as single nucleotide variants (SNVs), insertions/deletions (InDels), and short tandem repeats (STRs), can be identified through pipelines that integrate short read aligners for the mapping process and variant callers. Concerning the mapping process, BWA-MEM (11) is generally recommended due to its speed and accuracy in mapping DNA sequences to a large reference genome, such as the human genome (12,13). There is no single pipeline that optimally detects different types of variants. In fact, the combination of BWA-MEM (11) with Samtools (14) has demonstrated superior performance in SNP detection, while BWA-MEM (11) combined with GATK (15) has yielded better results for indels (16). In summary, the choice of pipeline should align with the specific research goal, and combining results from multiple pipelines can enhance overall outcomes (17).

In the field of variant identification, there are various approaches designed to detect specific sets of variants within genomic samples. One of the most widely utilized software tools for germline variant calling is the mapping tool HaplotypeCaller, part of the Genome Analysis Tool Kit (GATK-HC). GATK-HC identifies SNVs and InDels through haplotype construction, achieving a high level of accuracy at the expense of longer execution time. Considering the extensive array of tools available for the bioinformatic analysis of human genomic data and the imperative for precise diagnosis, we present a comprehensive benchmark of genome mapping and variant calling algorithms within a clinical-diagnostic framework. The benchmark aims to discern a fitting analysis workflow customized to the distinct requirements of laboratories primarily engaged in genetic medical diagnosis. It accomplishes this by undertaking a comparative evaluation of twenty-four distinct pipelines on two separate accessions of the same sample. The mapping software employed includes NGSEP, BWA, BWA-mem2, and Bowtie2, while variant caller software encompasses Deep Variant, Strelka, Free Bayes, NGSEP, Octopus, and GATK. For reference, the hg38 genome UCSC assembly was selected. Additionally, we assess the influence of sequencing on the variant calling process by analyzing two accessions of the same samples in two times (18). This approach is highly recommended as a best practice tool for variant identification in medical diagnosis (19). Strelka2 employs a Bayesian approach and represents continuous allelic frequencies while leveraging the expected genotype structure of normal samples to identify variants of potential interest (20).

Furthermore, DeepVariant, which is based on a deep convolutional neural network (15,20,21), has demonstrated high precision not only in SNV identification but also in the challenging task of detecting Copy Number Variants (CNVs). Other available tools for germline variation analysis include DRAGEN Bio-IT Platform, NGSEP, Octopus, and FreeBayes (22–25).

Considering the extensive array of tools available for the bioinformatic analysis of human genomic data and the imperative for precise diagnosis, we present a comprehensive benchmark of genome mapping and variant calling algorithms within a clinical-diagnostic framework. This benchmark is designed to discern a fitting analysis workflow customized to the distinct requirements of laboratories primarily engaged in genetic medical diagnosis. It accomplishes this by undertaking a comparative evaluation of twenty-four distinct pipelines on two separate accessions of the same sample. The mapping software employed includes NGSEP, BWA, BWA-mem2, and Bowtie2, while variant caller software encompasses Deep Variant, Strelka, Free Bayes, NGSEP, Octopus, and GATK. For reference, we selected the hg38 genome UCSC assembly. Additionally, we assess the influence of sequencing on the variant calling process by analyzing two accessions of the same samples.

## 3 Materials and methods

### 3.1 Data source

A systematic comparison of variant calling performance needs a gold-standard set of reference variant calls. To establish this standard, a FASTQ-formatted file from sample NA12872 was retrieved from the Sequence Read Archive Repository of the National Center for Biotechnology Information, provided by two distinct submitters: BGI-SHENZHEN submission sequenced with Illumina HiSeq 4000 (SRA ID: ERR1905890), and the original Broad Institute submission (SRA ID: SRR098401).

### 3.2 Mapping and variant calling

The process involved variant discovery and filtering through 24 distinct pipelines. Reads from both accessions were aligned against human reference genome, hg38 from the Broad Institute, using four different read aligners: BWA, BWA-MEM2, NGSEP, and Bowtie2, using default settings. BAM files were sorted to remove discrepancies, pairs with other orientations and pairs on different chromosomes. The evaluation included considerations for CPU consumption and runtime. The relevant reference genomes can be accessed at the following link: https://console.cloud.google.com/storage/browser/gcp-public-data--broad-references;tab=objects?prefix=&forceOnObjectsSortingFiltering=false.

The variant calling process was executed using six different tools: DeepVariant (15), Strelka2 (26), FreeBayes (25), NGSEP (23), Octopus (24), and GATK (27,28). To facilitate a clear comparison, default execution options were employed for each algorithm. After the process’s completion, 24 VCF files were obtained.

For the hg38 reference genome, we obtained the gold standard (release v. 4.2.1) from the GIAB FTP data repository (https://ftp-trace.ncbi.nlm.nih.gov/giab/ftp/release/). We compared each VCF file to the Gold Standard using the VCFGoldStandardComparator tool from NGSEP (23), utilizing Agilent SureSelect V.4.2.1 capture regions and Roche SeqCap (29) as references. File parsing was conducted using in-house scripts. All processes were executed on a DELL PowerEdge T550 Server with a 2 x Intel(R) Xeon(R) Gold 6338 CPU @ 2.00GHz, 128 CPUs, 256GB of RAM, and Ubuntu Server 20.04 LTS as the operating system.sing the Samtools sort utility (11). Summary metrics for this step included the percentage of mapped reads, percentage of properly paired reads, error rate, failed QC reads, MAPQ0 reads, discrepancies, pairs with other orientations, pairs on different chromosomes, CPU consumption, and runtime. The relevant reference genomes can be accessed at the following link: https://console.cloud.google.com/storage/browser/gcp-public-data--broad-references;tab=objects?prefix=&forceOnObjectsSortingFiltering=false.

## 4 Results

### 4.1 Mapping comparison

We evaluated metrics by comparing twenty-four pipelines resulting from the combination of 4 mapping and 6 Variant caller tools against the reference sample NA12878. The performance criteria were categorized into two main aspects: bioinformatic-quality assessment and computational behavior, encompassing resource consumption and execution time.

We evaluated two different accessions of the same samples: Broad Institute accession and BGI-SHENZHEN accession. To elucidate the distinctions in quality among the accessions, an assessment was conducted to evaluate the quality of the fasta files. The Broad Institute accession displayed substandard sequencing quality, as indicated by the Per base sequence quality statistics (Figure 1). None of the accessions exhibited the anticipated distribution of GC count, potentially accounting for the deviation in Ts/tv ratio. Furthermore, our analysis demonstrated compromised per base sequence quality in both samples, particularly evident in the Broad Institute accession (Figure 2). In this case, the mean quality surpassed the acceptable threshold for sequencing quality. Considering the Broad Institute accession’s esteemed status as a gold standard, its mean quality score should ideally surpass the accepted threshold for good quality sequencing. All the experiments were carried out with both accessions, to check the differences in the performance of the variant calling process in contrast with the quality of the sequencing process.

**Figure 1.**
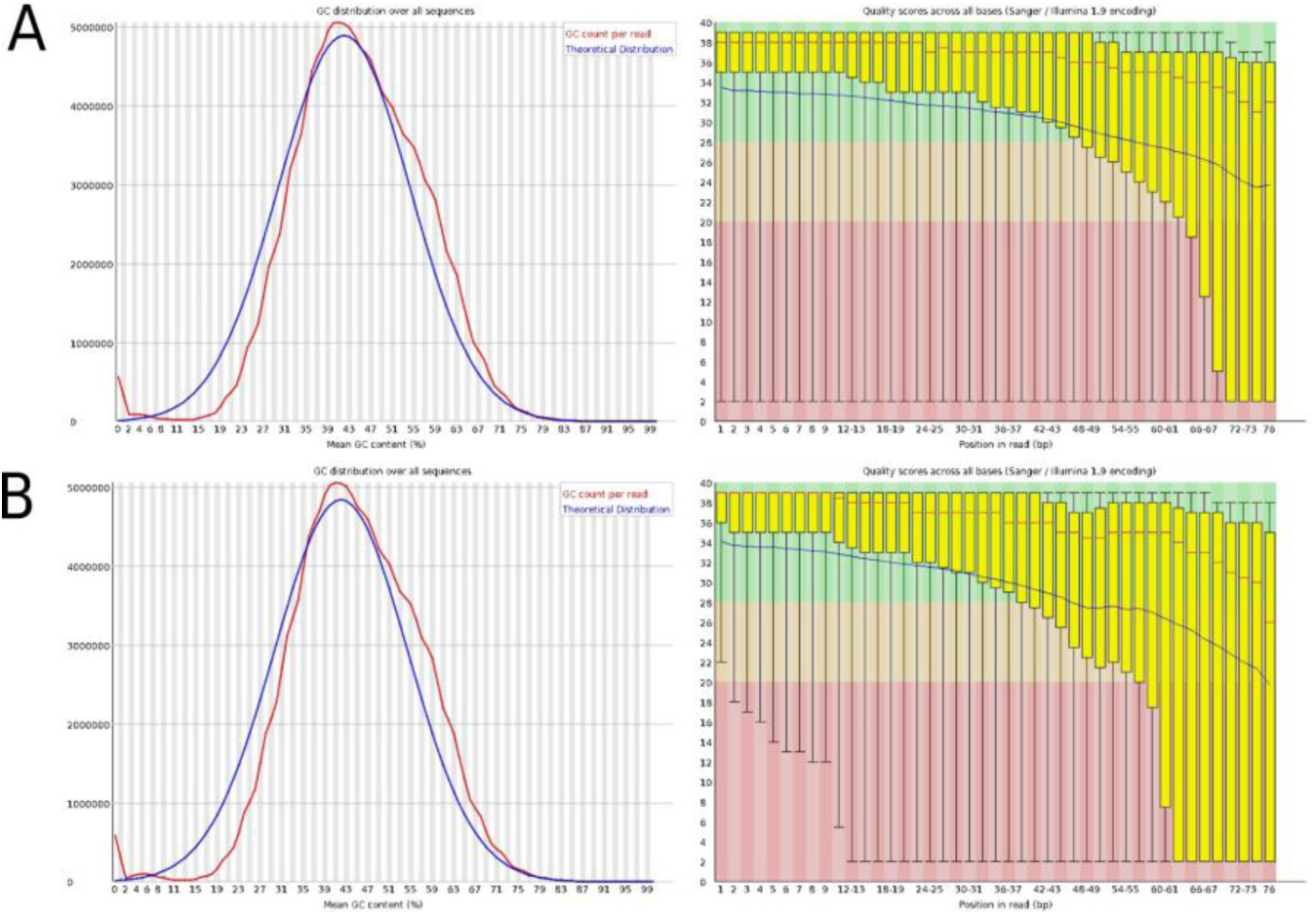
Fastq quality stats (per base sequence quality and per sequence GC content) for Broad Institute accession. A. Forward reads stats. B. Reverse reads stats.

**Figure 2.**
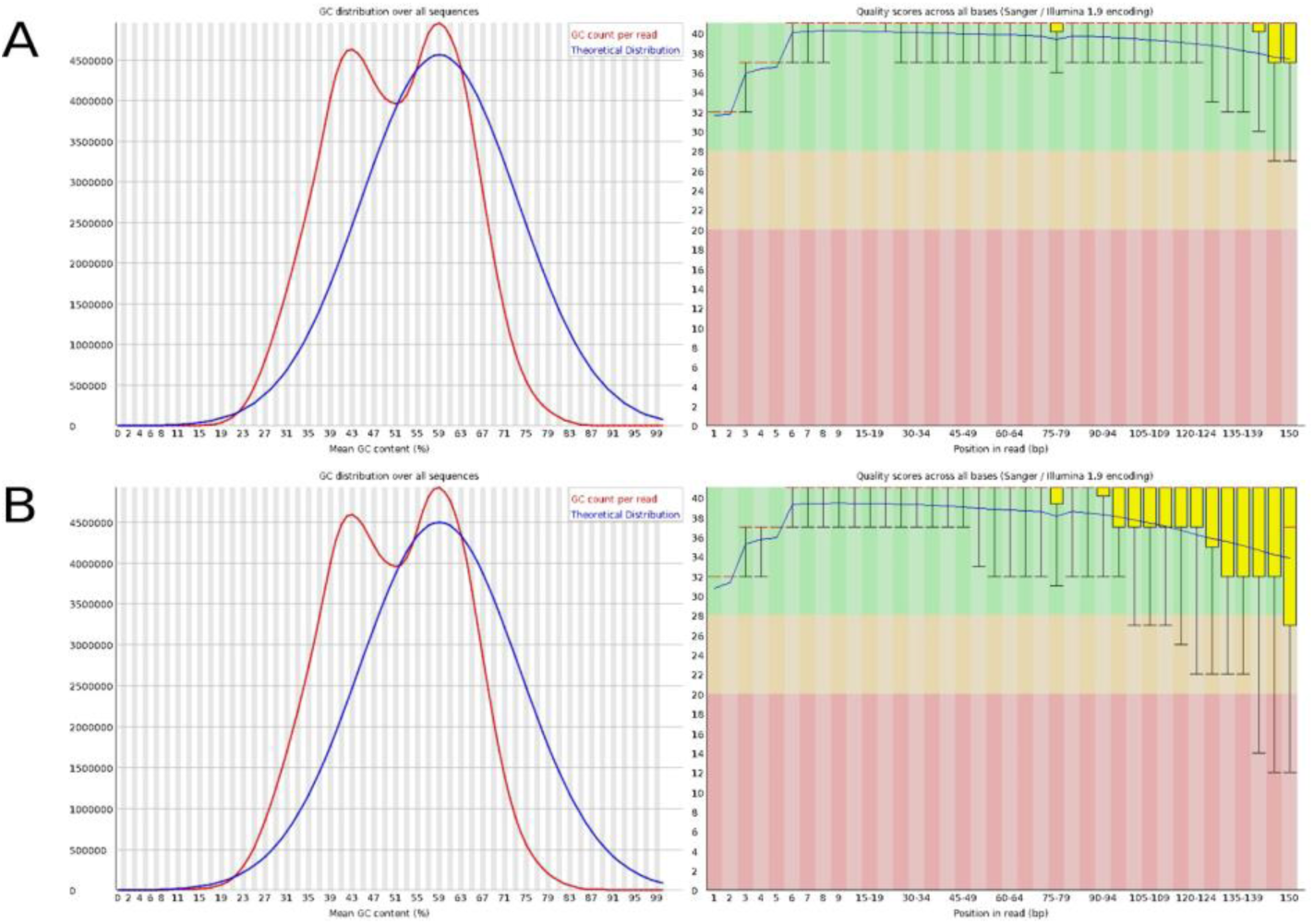
Fastq quality stats (per base sequence quality and per sequence GC content) for BGI-SHEZHEN accession. A. Forward reads stats. B. Reverse reads stats.

In our experiments, for both accessions NGSEP exhibited one of the lowest error rates, but it yielded the lowest percentage of properly paired reads mapped, amounting to less than 75%. Conversely, Bowtie2 achieved the highest mapping rates; however, it did exhibit a higher error rate in our experiments. As anticipated, both BWA and BWA-mem2 demonstrated striking similarity due to both employing the Burrows-Wheeler algorithm. They both achieved mapping rates exceeding 90% while maintaining the lowest error rates, as depicted in Figure 3. We aim to emphasize the variations in error rates between the same aligner used with two different accessions. For instance, in BWA-mem and BWA-mem2, differences in error rates are evident, as shown in Figure 3. To address this, we took into consideration key metrics such as execution time, CPU utilization (number of cores employed), and RAM usage (measured as the peak of resident memory set size). These metrics complement the aforementioned bioinformatic quality measures and underscore the algorithmic constraints inherent to each solution.

**Figure 3.**
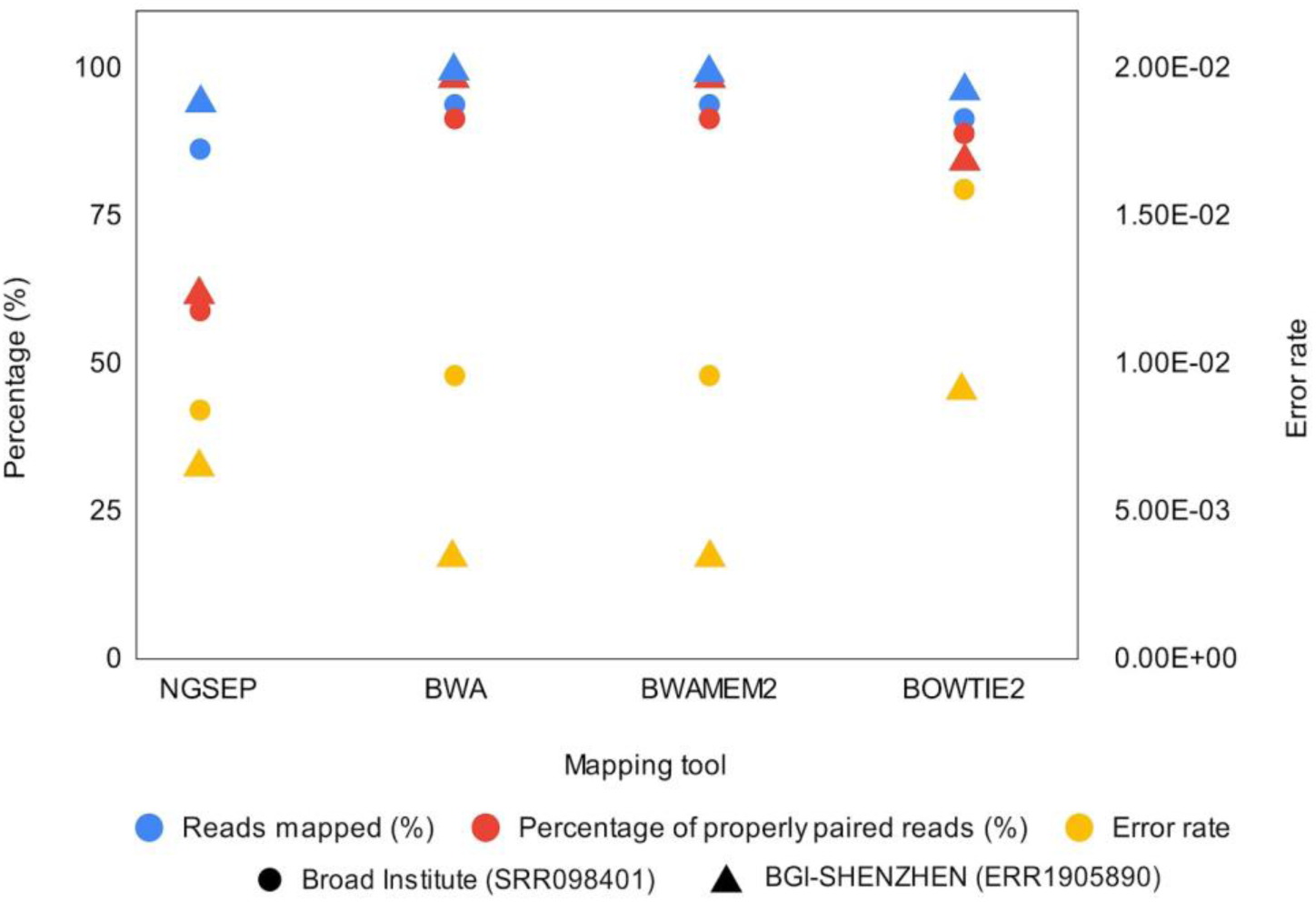
Percentage of reads mapped to the reference genome, properly paired reads and error rate in mapping process using NGSEP, BWA, BWA-mem2 and Bowtie2.

Among the evaluated software options, BWA-mem2 emerged as the fastest, boasting an average CPU usage when compared to other tools. However, it is noteworthy that BWA-mem2 exhibited a high resident set size, reaching a maximum of approximately 63GB. This behavior aligns with its algorithmic features, including optimizations for cache reuse, simplified algorithms, and the replacement of numerous small, fragmented memory allocations with fewer, larger contiguous ones. In contrast, both BWA-mem and Bowtie2 demonstrated similar execution times while allocating similar computational resources, as illustrated in Figure 4. On the other hand, NGSEP showcased an economical utilization of computational resources in terms of memory. Nevertheless, it is important to mention that its execution time exceeded 10 hours.

**Figure 4.**
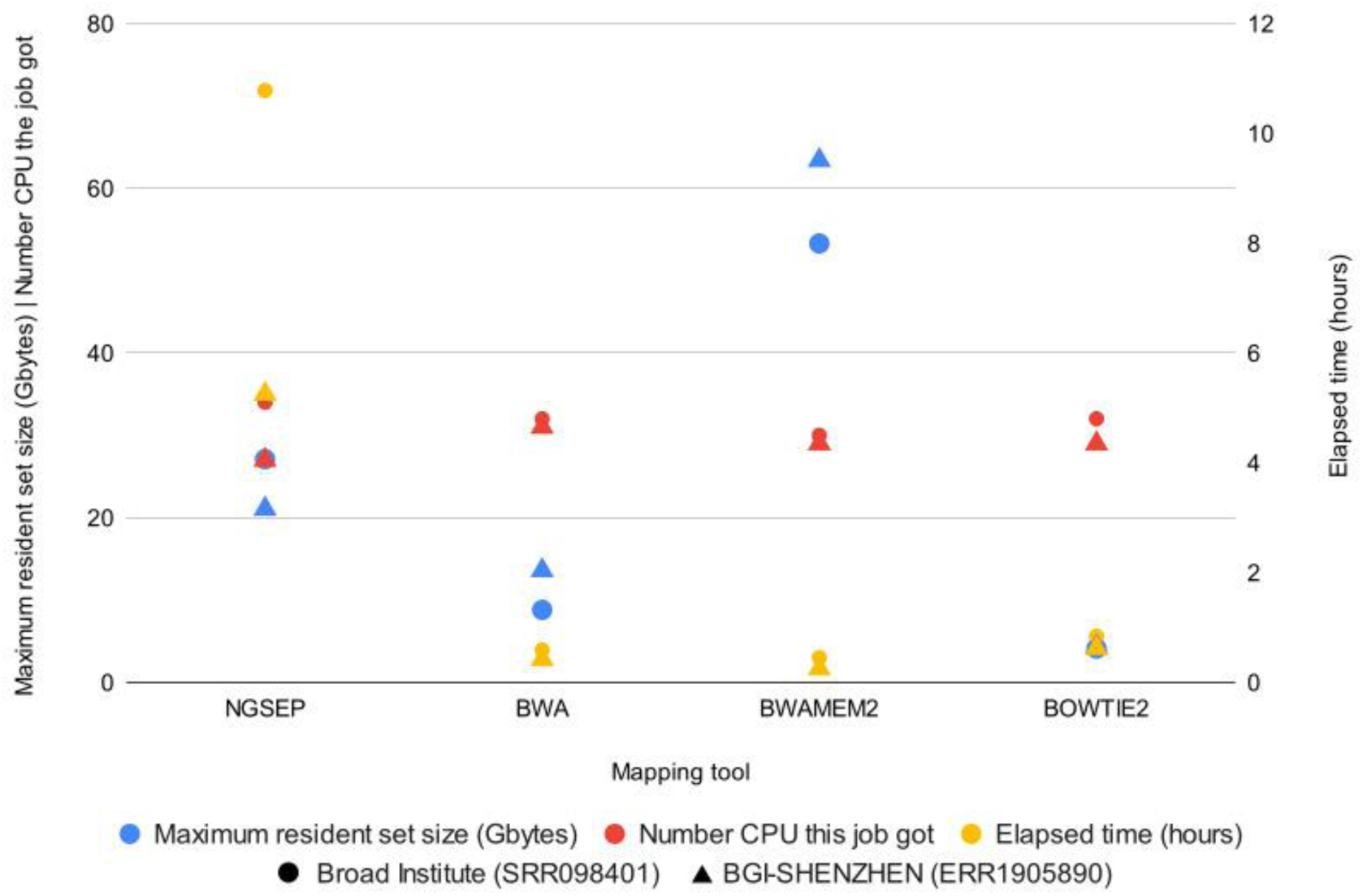
Computational resources used during mapping process using NGSEP, BWA, BWA-mem2 and Bowtie2.

### 4.2 Variant calling comparison

We conducted variant calling analysis employing six distinct variant caller software: Deep Variant, Free Bayes, Octopus, NGSEP, GATK, and Strelka. Assessment parameters included the count of SNPs, indels, and the transition/transversion ratio (ts/tv) identified by each caller. Among the pipelines, the NGSEP_NGSEP configuration, employing NGSEP as both the mapping tool and variant caller, demonstrated the highest SNP count at 22,106,766, followed by DeepVariant_Bowtie2 with 7,034,267 SNPs. In contrast, for BGI-SHEZHEN, the greatest SNP count reached 3,939,599 with Bowtie2_Deep Variant, trailed by NGSEP_NGSEP with 3,723,607 SNPs. These disparities in SNP counts persist across accessions. In reference to indels, the Broad Institute reference revealed a maximum of 709,383 indels detected (NGSEP_NGSEP), while the BGI-SHEZHEN reference showed 435,335 indels detected (Bowtie2_NGSEP). Both accessions demonstrated a low transition transversion ratio (ts/tv) ranging between 1 and 2.2, as depicted in Figure 5.

**Figure 5.**
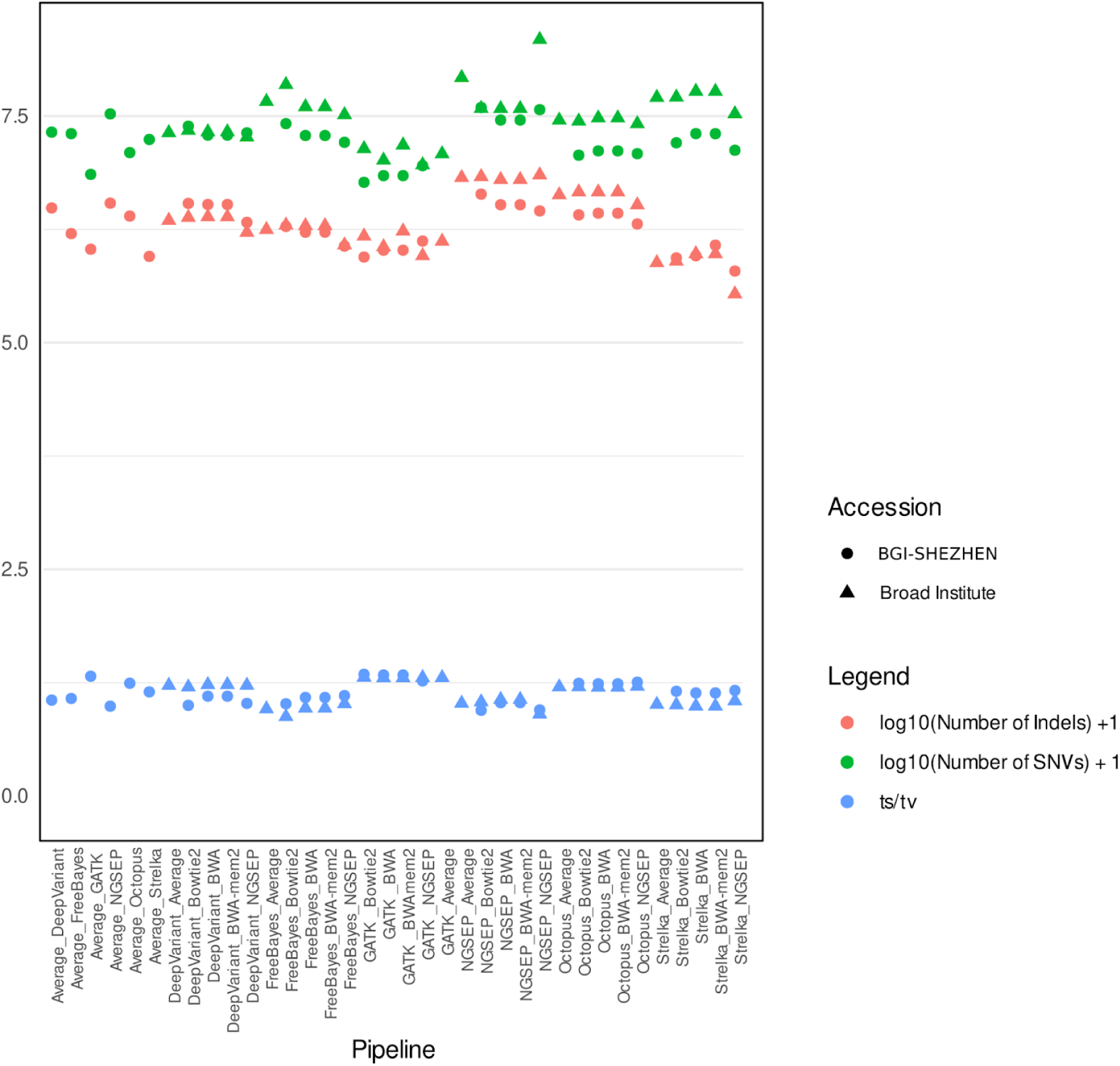
SNVs, Indels counts and ts/tv ratio for all variant calling pipelines evaluated with the two accessions.

We assessed computational performance among the mentioned variant calling software, examining elapsed time, maximum resident set size, and CPU utilization, mirroring our approach in the mapping comparison. NGSEP exhibited superior performance, as illustrated in Table 1, with an elapsed time of 3.82 hours. Following NGSEP were Free Bayes, Octopus, GATK, Strelka, and Deep Variant in terms of performance metrics.

**Table 1.**
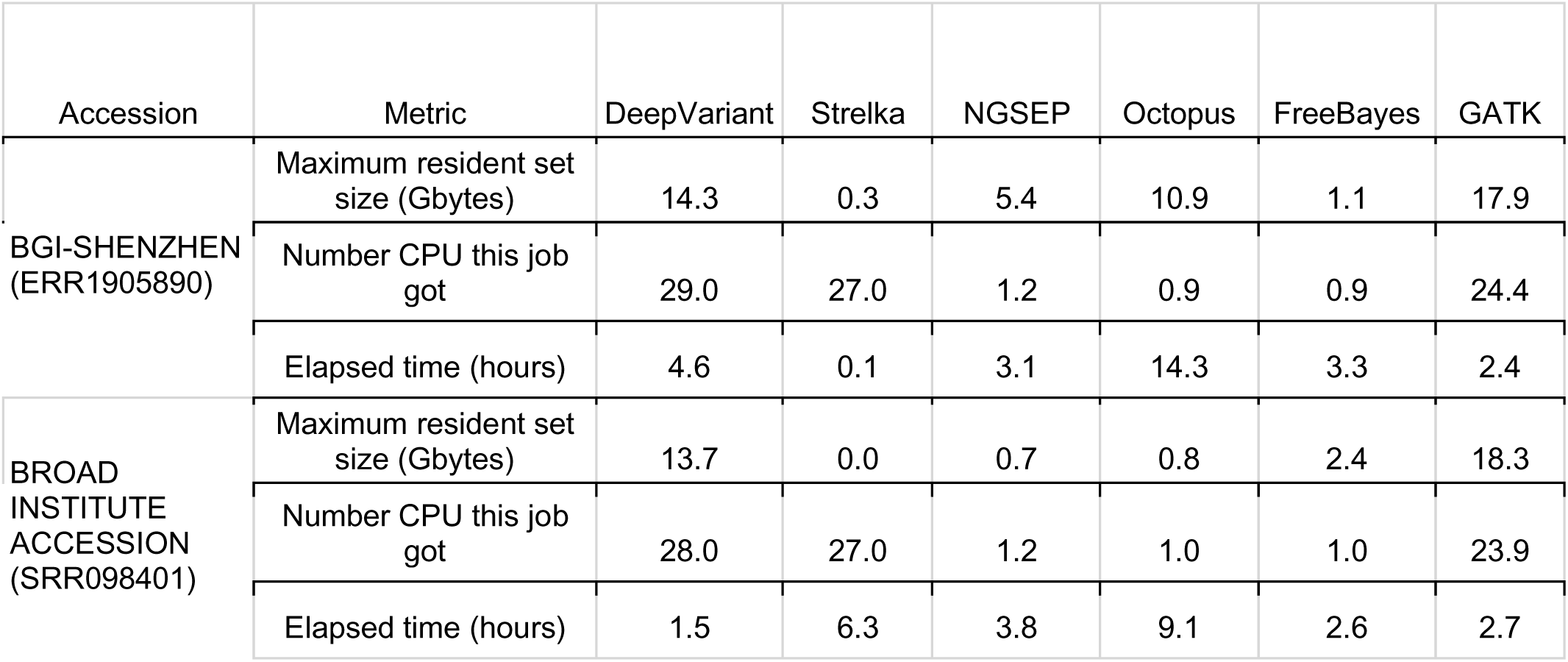
Computational performance for variant calling process.

| Accession | Metric | DeepVariant | Strelka | NGSEP | Octopus | FreeBayes | GATK |
| --- | --- | --- | --- | --- | --- | --- | --- |
| BGI-SHENZHEN<br>(ERR1905890) | Maximum resident set size (Gbytes) | 14.3 | 0.3 | 5.4 | 10.9 | 1.1 | 17.9 |
|  | Number CPU this job got | 29.0 | 27.0 | 1.2 | 0.9 | 0.9 | 24.4 |
|  | Elapsed time (hours) | 4.6 | 0.1 | 3.1 | 14.3 | 3.3 | 2.4 |
| BROAD<br>INSTITUTE<br>ACCESSION<br>(SRR098401) | Maximum resident set size (Gbytes) | 13.7 | 0.0 | 0.7 | 0.8 | 2.4 | 18.3 |
|  | Number CPU this job got | 28.0 | 27.0 | 1.2 | 1.0 | 1.0 | 23.9 |
|  | Elapsed time (hours) | 1.5 | 6.3 | 3.8 | 9.1 | 2.6 | 2.7 |

To assess the quality of variant calls, VCF files were compared against a gold standard reference over specific regions captured by two widely used commercial kits used in medical diagnosis — Sure Select V8 (SS) from Agilent and SeqCap (SC) from Illumina—.We assessed sensitivity as a measure of true positives for homozygous and heterozygous SNVs, Indels, and short tandem repeats (STRs) along the 24 different pipelines (4 mapping software + 6 variant calling software) in both accessions. Our findings underscore the substantial impact of the capture kit on these metrics, with SS demonstrating superior metrics compared to SC. As anticipated, homozygous variants outperformed heterozygous variants. Notably, a sensitivity difference of more than 10 percent was evident among accessions in heterozygous variants using the SS capture kit.

In the assessment of heterozygous variants, the most effective pipeline for SNVs was BWA-mem2_DeepVariant. For indels, the Broad Institute accession displayed superior metrics with BWA-mem2_Octopus, whereas the BGI-SHEZHEN accession yielded better results using BWA-mem2_DeepVariant. STRs typically pose significant challenges for mapping software. The top-performing results for both accessions were attained with BWA-mem2_GATK and BWA-mem2_Strelka2, demonstrating similar performance.

Additionally, our analysis across various pipelines revealed distinct outcomes, comparing homozygous variants that typically yield higher sensitivity metrics and fewer false positives (results not shown). The BGI-SHEZHEN accession delivered the optimal SNV result, achieving a sensitivity of 96.55% via the NGSEP_DeepVariant pipeline. Meanwhile, for this accession, BWA_Strelka2 yielded the highest sensitivity for indels (85.9%), and Bowtie_FreeBayes for STR (88.4%). In contrast, the Broad Institute accession showcased differing best-performing pipelines: BWA_NGSEP for SNVs (90.6%), BWA-mem2_Octopus for Indels (58.0%), and Bowtie_FreeBayes for STR (73.6%) (Figure 6).

**Figure 6.**
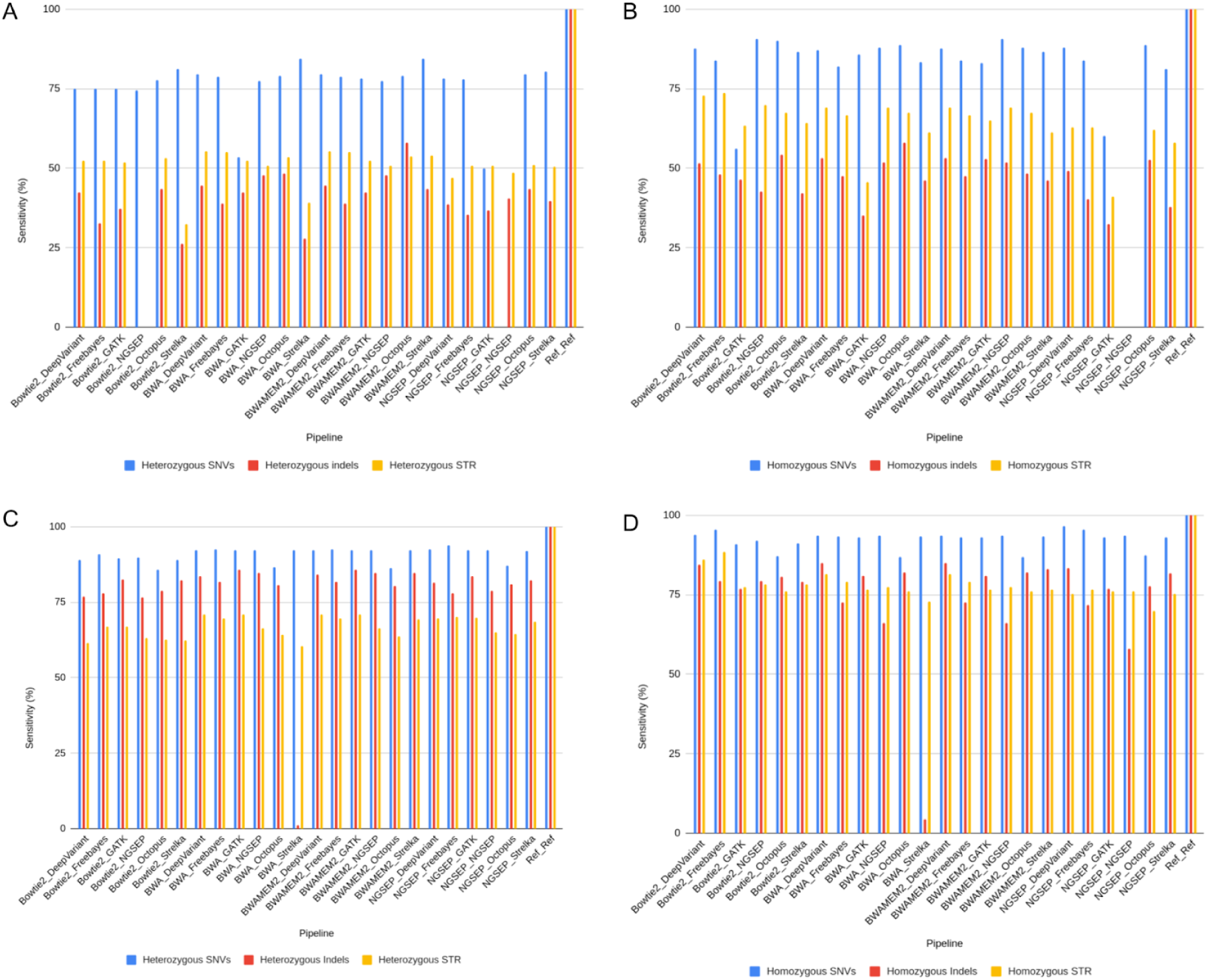
Sensitivity metrics for VCF gold standard comparison using SureSelect capture kit. A. Heterozygous variants for Broad Institute accession. B. Homozygous variants for Broad Institute accession. C. Heterozygous variants for BGI-SHENZHEN accession. D. Homozygous variants for BGI-SHENZHEN accession.

Another widely used capture kit in clinical diagnosis is SeqCap. By comparing previously acquired VCF data after filtering regions from this new capture kit, improvements were observed across all experiments. For heterozygous variants, sensitivities of 85.59% (Broad Institute) and 92.72% (BGI-SHEZHEN) were achieved for SNVs using the BWA_Strelka2 and NGSEP_FreeBayes pipelines, respectively. Regarding indels, sensitivities were 69.52% (Broad Institute accession) and 88.12% (BGI_SHEZHEN) using BWA_Octopus and BWA_GATK, respectively. Furthermore, STR detection sensitivity improved to 80.12% (Broad Institute) and 92.17% (BGI-SHEZHEN) utilizing Bowtie2_FreeBayes in both cases (Figure 7).

**Figure 7.**
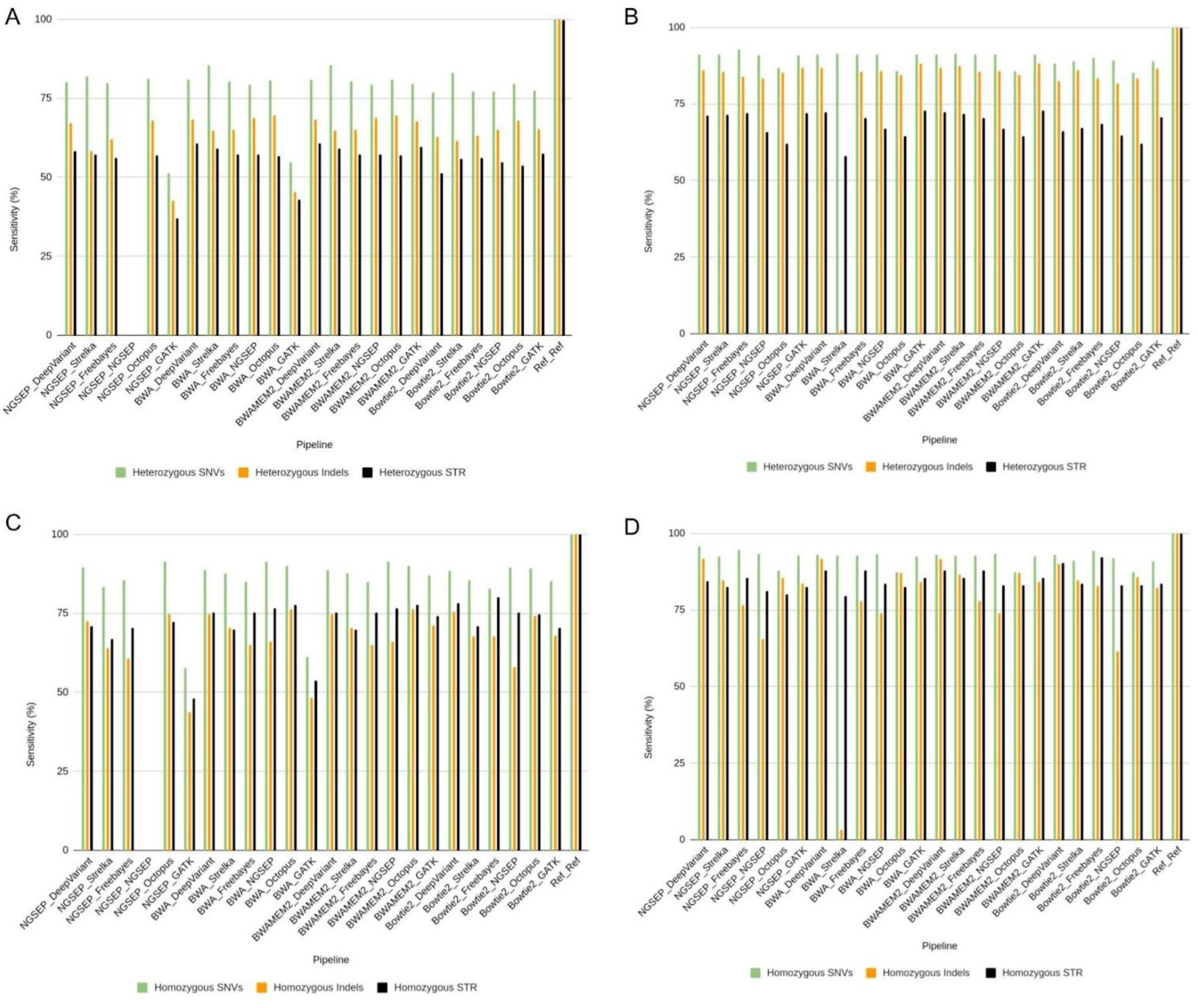
Sensitivity metrics for VCF gold standard comparison using SecCap capture kit. A. Heterozygous variants for Broad Institute accession. B. Homozygous variants for Broad Institute accession. C. Heterozygous variants for BGI-SHENZHEN accession. D. Homozygous variants for BGI-SHENZHEN accession.

The study yielded optimal metrics for homozygous variants using SC. Within the Broad Institute accession, sensitivity values were observed at 91.41% for SNVs, 76.35% for indels, and 80.12% for STR. Conversely, within the BGI-SHEZHEN accession, sensitivity registered at 95.73%, 91.74%, and 92.17% for the respective variants. The utilized pipelines for these metrics were NGSEP_Octopus and NGSEP_DeepVariant for SNVs, BWA_Octopus and BWA_DeepVariant for Indels, and Bowtie_FreeBayes for STR (Figure 7).

## 5 Discussion

In the realm of clinical diagnostics, the establishment of standardized pipelines for exome analysis is imperative due to the pervasive inconsistencies affecting variant identification, annotation, filtration, classification, and reporting. The discrepancy in variant classifications among submitters, as evidenced by the 17% variability in ClinVar variants, underscores the urgency for uniform guidelines in interpreting variants (30). The complexity of genome-wide variant analysis amplifies the challenge for clinical laboratories, necessitating a structured approach from raw data processing to phenotype-associated variant reporting. The bioinformatics pipeline involves a multitude of steps, encompassing diverse algorithms, software, databases, and operational environments. Yet, the absence of a defined standard for integrating these components to analyze outputs across sequencing platforms impedes the reliability and comparability of results, hampering their effective use in patient care (17).

Therefore, establishing standardized pipelines is therefore indispensable to enhance the consistency, accuracy, and clinical utility of exome analysis in healthcare settings and optimizing computational resources is crucial to minimize processing time and maximize throughput.

Mapping is a fundamental process in variant detection in clinics. Borrows-Wheeler transform was developed in 1994 and it started to call bioinformatics attention around 2010, Bowtie, BWA implemented this as the basis of their mapping software (11,31). BWA was originally developed by Heng Li (11). It constructs a prefix based on a transformation, it is optimized for short reads, especially single end but it also supports paired ends. In the original publication of BWA, it accomplished a fastest execution time compared with Bowtie2 and a higher percentage of paired reads. Despite the use of the same algorithm, we evidence differences in the use of computational resources between Bowtie, BWA-mem and BWA-mem2, a previous study showed that low quality samples can change the performance among different software, additionally, they highlight that all repetitive regions are usually aligned incorrectly decreasing accuracy (32).

We also evidenced difference in metrics between capture kits despite both are based on hybridization and sonication for fragmenting DNA sample, one of the main differences is that SureSelect uses RNA probes, while SeqCap uses DNA with a bigger size per library (33). Moreover, manufacturer specifies an alignment percentage of 30-70% for SureSelect and 70-80% for SeqCap, which can explain why the analysis with SeqCap showed a better performance, this capture kit is the one with a better mapping rate among capture kits evaluated by Samorodnitsky, *et al.,* 2015 (SureSelect, HaloPlex, Nextera and SeqCap) (33).

Regarding the inconsistent in ts/tv ratio, some studies have reported the association between ancestry and variants, a value of 2.0 in whole genome sequencing has been observed in African ancestry population. European, Asian, and American samples usually show lower values (34), NA12878 sample has been described as European Ancestry, our observed ts/tv ratios were lower in both accession than the reported for exome data, that is usually between 2.6 and 3 (34) which may be explained by the capture kit used that is taking intron sequences deviating the ratio, or the low-quality sequencing.

In our experiments, the best mapping tool was BWA due to its computing performance and mapping quality, using less than one hour and the lowest error rate. Some studies have shown the good performance of this mapping software in SNV and indel detection using samtools as a variant calling tool (14,16), however it has shown that it has a weak performance with heterozygous SNVs (16).

We evidenced the influence of the mapping process to variant calling sensitivity. The sequence quality showed to be fundamental in the variant detection, with the BGI-SHEZHEN accession we were able to obtain sensitivity values up to 93%, meanwhile with a lower quality sample the maximum sensitivity was 85%. It is also important to select the variant type of interest (SNVs, Indels, STR) for optimizing the pipeline.

In concordance with prior research findings, DeepVariant has demonstrated superior performance. Previous studies have documented that DeepVariant, while making fewer calls, exhibits a low rate of false positives and fewer false negatives in identifying variants of interest (21). Consequently, there have been suggestions for its widespread adoption on a larger scale, with some reports even indicating a reduction in computational costs (35). However, in our specific case, this tool has exhibited the highest computational resource consumption. This limitation restricts its extensive use for medical diagnoses and the analysis of large sample cohorts.

In short-read sequencing, one of the primary challenges arises in the presence of tandem repeated regions, primarily due to the inadequate sequencing method’s representation of these regions.

Generally, variant calling algorithms exhibit reduced performance in identifying indels compared to SNVs, making the identification of these variants a significant challenge (24). Nevertheless, as observed in other studies, our data indicated that Octopus outperformed in the identification of indels. It has been reported that this tool has the capability to detect a broader range of indels and demonstrates greater precision when identifying deletions smaller than 15 base pairs (24). Furthermore, it exhibited a better performance when used in conjunction with BWA for mapping (9), a finding that aligns with our own data.

In consequence, the optimal pipeline for the SureSelect capture kit involves using BWA-mem2 for mapping and DeepVariant for variant detection. Despite not yielding the best results in all tests, this combination demonstrated superior performance for SNVs, processing within 8 hours, and achieving sensitivity values exceeding 90% when assessed using BGI-SHEZHEN data.

In situations of limited computational resources, the BWA-mem2_Strelka2 pipeline offers an efficient solution, significantly reducing processing time to a few seconds while achieving a sensitivity of 92.21% based on BGI-SHEZHEN accession data. This accelerated processing time holds considerable promise for expediting clinical recurrent tasks, where minimizing opportunity time is crucial. Noticeable variations in metrics were observed between accessions, with lower metrics recorded in the Broad Institute dataset, possibly due to differences in sequencing processing methodologies. The BGI-SHEZHEN accession utilized the SeqCap V5 kit, while information regarding the lab processing for the Broad Institute accession remains unavailable.

Bioinformatic tools have shown a relevant importance in the past years in clinical context, pipeline selection can improve a patient’s diagnosis (36). Considering that the current guidelines of ACMG (37) are designed to classify SNV and there is more information about the functional effect of SNV than other types of variation, the most accurate and fast pipeline was BWA and Strelka2.

We obtained better metrics for the SecCap capture kit than for the SureSelect V8. Considering the best metrics in both kits, the best workflow for SNVs were BWA, BWA-mem2, and NGSEP in combination with DeepVariant and Strelka2. Regarding computational resource consumption, the most sensitive workflow with a smaller number of false positives was the BWA_Strelka2 workflow.

On the other hand, if the main purpose is to detect indels properly, the best solutions are BWA and BWA-mem2 as mapping algorithms, along with Octopus, GATK, and DeepVariant as variant callers. Where the fastest and least resource-consuming option is BWA_Octopus. Meanwhile, for STR, the best pipeline in most of the experiments was Bowtie2_FreeBayes.

Lastly, it is crucial to emphasize that, when selecting the most appropriate tool for genetic diagnosis, consideration must be given to computational resources. Various tools have demonstrated their efficacy in mapping and variant calling for clinical purposes. Laboratories equipped with GPUs or access to cloud processing may find tools like DeepVariant advantageous for variant identification, owing to their ability to handle computational-intensive tasks. Conversely, in scenarios involving large-scale processing with limited computational resources, tools such as BWA_Strelka2 offer significant benefits.

## Data Availability

All data produced in the present study are available upon reasonable request to the authors

## Contributions by author

Johanna Stepanian: Conceptualization, Data curation, Formal analysis, Investigation, Methodology, Validation, Visualization, Writing - original draft, Writing - review and editing.

Diego Saldaña: Conceptualization, Data curation, Investigation, Methodology, Project administration, Supervision, Validation, Visualization, Writing - original draft, Writing - review and editing.

Daniel Mahecha: Conceptualization, Data curation, Investigation, Methodology, Software, Supervision, Validation, Visualization, Writing - original draft, Writing - review and editing. Laura Mazabel: Data curation, Data analysis, Investigation.

Jorge Iván Díaz-Riaño: Conceptualization, Investigation, Methodology, Project administration, Resources, Software, Supervision, Validation, Visualization, Writing - original draft, Writing - review and editing.

## 6 Acknowledgments

We want to thank Dr. Silvia Maradei for helpful suggestions and review of the manuscript.

## 9 References

1. Koboldt DC. Best practices for variant calling in clinical sequencing [Internet]. Vol. 12, Genome Medicine. Springer Science and Business Media LLC; 2020. Available from: 10.1186/s13073-020-00791-w

2. van Dijk EL, Auger H, Jaszczyszyn Y, Thermes C. Ten years of next-generation sequencing technology [Internet]. Vol. 30, Trends in Genetics. Elsevier BV; 2014. p. 418–26. Available from: 10.1016/j.tig.2014.07.001

3. Pereira PCB, Melo FM, De Marco LAC, Oliveira EA, Miranda DM, Simões e Silva AC. Whole-exome sequencing as a diagnostic tool for distal renal tubular acidosis [Internet]. Vol. 91, Jornal de Pediatria. Elsevier BV; 2015. p. 583–9. Available from: 10.1016/j.jped.2015.02.002

4. Renkema KY, Stokman MF, Giles RH, Knoers NVAM. Next-generation sequencing for research and diagnostics in kidney disease [Internet]. Vol. 10, Nature Reviews Nephrology. Springer Science and Business Media LLC; 2014. p. 433–44. Available from: 10.1038/nrneph.2014.95.

5. Krøigård AB, Thomassen M, Lænkholm AV, Kruse TA, Larsen MJ. Evaluation of Nine Somatic Variant Callers for Detection of Somatic Mutations in Exome and Targeted Deep Sequencing Data [Internet]. Jordan IK, editor. Vol. 11, PLOS ONE. Public Library of Science (PLoS); 2016. p. e0151664. Available from: 10.1371/journal.pone.0151664

6. McCombie WR, McPherson JD, Mardis ER. Next-Generation Sequencing Technologies [Internet]. Vol. 9, Cold Spring Harbor Perspectives in Medicine. Cold Spring Harbor Laboratory; 2018. p. a036798. Available from: 10.1101/cshperspect.a036798

7. Karczewski KJ, Francioli LC, Tiao G, Cummings BB, Alföldi J, Wang Q, et al. The mutational constraint spectrum quantified from variation in 141,456 humans [Internet]. Vol. 581, Nature. Springer Science and Business Media LLC; 2020. p. 434–43. Available from: 10.1038/s41586-020-2308-7.

8. Bycroft C, Freeman C, Petkova D, Band G, Elliott LT, Sharp K, et al. The UK Biobank resource with deep phenotyping and genomic data. Vol. 562, Nature. Springer Science and Business Media LLC; 2018. p. 203–9. Available from: 10.1038/s41586-018-0579-z.

9. Barbitoff YA, Abasov R, Tvorogova VE, Glotov AS, Predeus AV. Systematic benchmark of state-of-the-art variant calling pipelines identifies major factors affecting accuracy of coding sequence variant discovery [Internet]. Vol. 23, BMC Genomics. Springer Science and Business Media LLC; 2022. Available from: 10.1186/s12864-022-08365-3.

10. American Society of Human Genetics Board of Directors and American College of Medical Genetics Board of Directors. Points to Consider: Ethical, Legal, and Psychosocial Implications of Genetic Testing in Children and Adolescents. Am J Hum Genet. November 1995;57(5):1233–41.

11. Li H. Aligning sequence reads, clone sequences and assembly contigs with BWA-MEM. 2014.

12. Jung Y, Han D. BWA-MEME: BWA-MEM emulated with a machine learning approach. Bioinformatics. April 2022;38(9):2404–13.

13. Liu S, Wang Y, Wang F. A fast read alignment method based on seed-and-vote for next generation sequencing. BMC Bioinformatics. December 2016;17(17):466.

14. Danecek P, Bonfield JK, Liddle J, Marshall J, Ohan V, Pollard MO, et al. Twelve years of SAMtools and BCFtools. GigaScience. February 2021;10(2):giab008.

15. Poplin R, Chang PC, Alexander D, Schwartz S, Colthurst T, Ku A, et al. A universal SNP and small-indel variant caller using deep neural networks. Nat Biotechnol. November 2018;36(10):983–7.

16. Hwang S, Kim E, Lee I, Marcotte EM. Systematic comparison of variant calling pipelines using gold standard personal exome variants [Internet]. Vol. 5, Scientific Reports. Springer Science and Business Media LLC; 2015. Available from: 10.1038/srep17875.

17. Brownstein CA, Beggs AH, Homer N, Merriman B, Yu TW, Flannery KC, et al. An international effort towards developing standards for best practices in analysis, interpretation and reporting of clinical genome sequencing results in the CLARITY Challenge [Internet]. Vol. 15, Genome Biology. Springer Science and Business Media LLC; 2014. p. R53. Available from: 10.1186/gb-2014-15-3-r53.

18. Ren S, Bertels K, Al-Ars Z. Efficient Acceleration of the Pair-HMMs Forward Algorithm for GATK HaplotypeCaller on Graphics Processing Units. Evol Bioinforma Online. March 2018;14:1176934318760543.

19. Van der Auwera GA, Carneiro MO, Hartl C, Poplin R, del Angel G, Levy-Moonshine A, et al. From FastQ data to high confidence variant calls: the Genome Analysis Toolkit best practices pipeline. Curr Protoc Bioinforma Ed Board Andreas Baxevanis Al. October 2013;11(1110):11.10.1–11.10.33.

20. Pei S, Liu T, Ren X, Li W, Chen C, Xie Z. Benchmarking variant callers in next-generation and third-generation sequencing analysis. Brief Bioinform. May 2021;22(3):bbaa148.

21. Lin YL, Chang PC, Hsu C, Hung MZ, Chien YH, Hwu WL, et al. Comparison of GATK and DeepVariant by trio sequencing. Sci Rep. February 2022;12(1):1809.

22. Goyal A, Kwon HJ, Lee K, Garg R, Yun SY, Kim YH, et al. Ultra-Fast Next Generation Human Genome Sequencing Data Processing Using DRAGENTM Bio-IT Processor for Precision Medicine. Open J Genet. February 2017;7(1):9–19.

23. Tello D, Gil J, Loaiza CD, Riascos JJ, Cardozo N, Duitama J. NGSEP3: accurate variant calling across species and sequencing protocols. Schwartz R, editor. Vol. 35, Bioinformatics. Oxford University Press (OUP); 2019. p. 4716–23. Available from: 10.1093/bioinformatics/btz275.

24. Cooke DP, Wedge DC, Lunter G. A unified haplotype-based method for accurate and comprehensive variant calling. Nat Biotechnol. July 2021;39(7):885–92.

25. Garrison E, Marth G. Haplotype-based variant detection from short-read sequencing [Internet]. arXiv; 2012 [cited May 19 2024]. Available at: http://arxiv.org/abs/1207.3907

26. Kim S, Scheffler K, Halpern AL, Bekritsky MA, Noh E, Källberg M, et al. Strelka2: fast and accurate calling of germline and somatic variants. Nat Methods. August 2018;15(8):591–4.

27. DePristo MA, Banks E, Poplin R, Garimella KV, Maguire JR, Hartl C, et al. A framework for variation discovery and genotyping using next-generation DNA sequencing data. Vol. 43, Nature Genetics. Springer Science and Business Media LLC; 2011. p. 491–8. Available from: 10.1038/ng.806

28. McKenna A, Hanna M, Banks E, Sivachenko A, Cibulskis K, Kernytsky A, et al. The Genome Analysis Toolkit: A MapReduce framework for analyzing next-generation DNA sequencing data. Genome Res. September 2010;20(9):1297–303.

29. Chen R, Im H, Snyder M. Whole-Exome Enrichment with the Roche NimbleGen SeqCap EZ Exome Library SR Platform. Cold Spring Harb Protoc. March 2015;2015(7):pdb.prot084855.

30. Zhang K, Lin G, Han D, Han Y, Wang J, Shen Y, et al. An Initial Survey of the Performances of Exome Variant Analysis and Clinical Reporting Among Diagnostic Laboratories in China [Internet]. Vol. 11, Frontiers in Genetics. Frontiers Media SA; 2020. Available from: 10.3389/fgene.2020.582637

31. Ferragina P, Giancarlo R, Manzini G, Sciortino M. Boosting textual compression in optimal linear time. J ACM. July 2005;52:688–713.

32. Yu X, Guda K, Willis J, Veigl M, Wang Z, Markowitz S, et al. How do alignment programs perform on sequencing data with varying qualities and from repetitive regions? [Internet]. Vol. 5, BioData Mining. Springer Science and Business Media LLC; 2012. Available from: 10.1186/1756-0381-5-6.

33. Samorodnitsky E, Datta J, Jewell BM, Hagopian R, Miya J, Wing MR, et al. Comparison of Custom Capture for Targeted Next-Generation DNA Sequencing. J Mol Diagn JMD. January 2015;17(1):64–75.

34. Wang J, Raskin L, Samuels DC, Shyr Y, Guo Y. Genome measures used for quality control are dependent on gene function and ancestry. Bioinformatics. February 2015;31(3):318–23.

35. Yun T, Li H, Chang PC, Lin MF, Carroll A, McLean CY. Accurate, scalable cohort variant calls using DeepVariant and GLnexus. Bioinformatics. April 2021;36(24):5582–9.

36. Cowley MJ, Liu YC, Oliver KL, Carvill G, Myers CT, Gayevskiy V, et al. Reanalysis and optimisation of bioinformatic pipelines is critical for mutation detection. Hum Mutat. April 2019;40(4):374–9.

37. Richards S, Aziz N, Bale S, Bick D, Das S, Gastier-Foster J, et al. Standards and guidelines for the interpretation of sequence variants: a joint consensus recommendation of the American College of Medical Genetics and Genomics and the Association for Molecular Pathology [Internet]. Vol. 17, Genetics in Medicine. Elsevier BV; 2015. p. 405–24. Available from: 10.1038/gim.2015.30

